# Association of olfactory dysfunction with hospitalization for COVID-19: a multicenter study in Kurdistan

**DOI:** 10.1101/2020.07.26.20158550

**Authors:** Hosna Zobairy, Erfan Shamsoddin, Mohammad Aziz Rasouli, Nasrollah Veisi Khodlan, Ghobad Moradi, Bushra Zareie, Sara Teymori, Jalal Asadi, Ahmad Sofi-Mahmudi, Ahmad R Sedaghat

## Abstract

**Objective:** To evaluate the association of olfactory dysfunction (OD) with hospitalization for COVID-19.

**Study Design:** Multi-center cohort study.

**Setting:** Emergency departments of thirteen COVID-19-designed hospitals in Kurdistan province, Iran.

**Subjects and Methods:** Patients presenting with flu-like symptoms who tested positive by RT-PCR for COVID-19 between May 1^st^ and 31^st^, 2020. At the time of presentation and enrollment, patients were asked about the presence of OD, fever, cough, shortness of breath, headache, rhinorrhea and sore throat. The severity of OD was assessed on an 11-point scale from 0 (none) to 10 (anosmia). Patients were either hospitalized or sent home for outpatient care based on standardized criteria.

**Results:** Of 203 patients, who presented at a mean of 6 days into the COVID-19 disease course, 25 patients (12.3%) had new OD and 138 patients (68.0%) were admitted for their COVID-19. Patients admitted for COVID-19 had a higher prevalence of all symptoms assessed, including OD (p<0.05 in all cases), and OD identified admitted patients with 84.0% sensitivity and 34.3% specificity. On univariate logistic regression, hospitalization was associated with OD (odds ratio [OR] = 2.47, 95%CI: 1.085–6.911, p=0.049). However, hospitalization for COVID-19 was not associated with OD (OR=3.22, 95% CI: 0.57–18.31, p=0.188) after controlling for confounding demographics and comorbidities.

**Conclusion:** OD may be associated with hospitalization for (and therefore more severe) COVID-19. However, this association between OD and COVID-19 severity is more likely driven by patient characteristics linked to OD, such as greater numbers of COVID-19 symptoms experienced or high-risk comorbidities.

## Introduction

The 2019 Coronavirus Disease (COVID-19) is a pandemic that continues to spread and pose an international public health emergency since it was first identified in December 2019.^1, 2^ With every passing day, the scientific community continues to learn more about the disease and, in fact, COVID-19-related symptoms continue to be recognized and described. While COVID-19 was initially characterized by symptoms of fever, cough, muscle aches and/or fatigue, dyspnea, headache, sore throat, and gastrointestinal symptoms,^3^ olfactory dysfunction (OD) has been reported to be a common symptom of COVID-19.^4, 5^ In fact, at this time, OD is well-accepted as a symptom of COVID-19, occurring in approximately 50% of COVID-19 positive individuals.^4, 5^

OD has been highlighted as an early symptom of COVID-19,^6^ reportedly to occurring at a mean of 3-4 days into the COVID-19 disease course.^7, 8^ OD may be the first symptom of COVID-19 in up to a quarter of COVID-19 patients.^9, 10^ As such, OD has been viewed as an early marker of COVID-19 that may be helpful in the early diagnosis of COVID-19, which may consequently reduce spread of the disease.^9, 11-13^ OD has also been proposed to be a prognostic indicator of COVID-19 disease severity. However, there have conflicting reports as to whether OD portends a milder or more severe COVID-19 disease course.^7, 14^

The identification of prognostic indicators for the COVID-19 disease course may be important for several reasons. Perhaps most importantly, patients anticipated to have the greatest disease severity could be identified early and treated aggressively. The recent discovery of remdesivir as an antiviral medication for COVID-19 has been an exciting development in the treatment of COVID-19.^15^ Other potential antiviral medications for COVID-19 are in clinical trials as well.^16^ However, because of the likely scarcity in available doses—which has already been observed for remdesivir—and the likely need to initiate antiviral treatment early in the disease course, predictors of a severe COVID-19 disease course are needed. Herein, we report the results of a multicenter study of COVID-19 patients who presented to 13 hospitals across the Kurdistan province of Iran. The objective of our study was to determine the association of OD with disease severity in COVID-19 patients, with hospitalization used as the metric for severe disease.

## Methods

### Subjects, setting and design

This study was conducted from May 1^st^ to 31^st^ 2020 at 13 distinct hospitals in Kurdistan province of Iran. We used convenience sampling to recruit patients presenting for flu-like symptoms to emergency departments at participating hospitals. All patients were the residents of Kurdistan province. The study protocol was approved by the Ethics Committee of Kurdistan University of Medical Sciences (ref. no. IR.MUK.REC.1399.006). All participants provided informed consent for inclusion in this study.

The inclusion criteria for our study included laboratory-confirmed COVID-19 infection (reverse transcription-polymerase chain reaction [RT-PCR] from nasopharyngeal swabs), age greater than 18 years, and the ability to complete the study questionnaire. Exclusion criteria included patients who had a history of OD for more than 1 month before presentation.

Patient were directly recruited from hospitals which were specially designated for treatment of COVID-19 patients in Kurdistan province, Iran. Once the patients visited the emergency department of one of these hospitals, they were evaluated and managed based on the contemporary COVID-19 protocol of Iran.^17^ At that point, they were also tested for COVID-19 by RT-PCR. Patients were deemed to require hospitalization if they presented with severe shortness of breath, were prescribe to receive non-invasive oxygen therapy, had SpO2<93% or respiratory rate>30.^17^ Patients were followed through the end of their COVID-19 course for admission to an intensive care unit (ICU), re-hospitalization, death, or symptomatic resolution of the disease.

### Study questionnaire

The questionnaire consisted of three parts: demographic information, medical history, and COVID-19-related symptomatology. Demographic information collected included age, gender, and history of active smoking. Medical history that was assessed included a history of chronic rhinosinusitis and allergic rhinitis. A general medical history was taken specifically to assess for a history of diabetes mellitus, hypertension, heart disease (coronary artery disease or congestive heart failure), cancer, and respiratory disease (asthma or chronic obstructive pulmonary disease).

The COVID-19 symptoms that were assessed included the presence of cough, fever, shortness of breath, sore throat, rhinorrhea and headaches at the time of presentation (as a binary variable: yes/no). OD at the time of presentation was assessed as a binary variable (yes/no) and its severity on an 11-point Likert scale ranging from 0 (no OD) to 10 (complete anosmia). The total number of days with symptoms attributable to COVID-19 was also assessed at the time of presentation.

### Statistical analysis

We used Statistical Package for the Social Sciences for Windows (SPSS version 25,0; IBM Corp, Armonk, NY, USA) to perform the statistical analyses. The potential associations between epidemiological, clinical and olfactory outcomes have been assessed through cross-tab generation between two variables (binary or categorical variables) and Chi-square test. We used logistic and linear regression to fit a model for anosmia outcomes. Incomplete responses were excluded from the analysis. A level of p<0.05 was used to determine statistical significance.

## Results

### Study participants

A total of 203 individuals participated in this study. The mean age of the participants was 49.2 years, with standard deviation (SD) of 16.6 years (range 20-114 years). There were 91 females (44.8%) and 112 males (55.2%). These study participants presented at a mean 6.4 days (SD: 4.2 days) after the onset of COVID-19 symptoms. Of the entire cohort, 138 were hospitalized while 65 were not hospitalized. The hospitalized patients presented at a mean 6.7 days (SD: 4.3) into the COVID-19 course and were admitted to the hospital for a mean of 5.2 days (range 1 – 25). None of the hospitalized patients required transfer to an ICU or were re-admitted for COVID-19. The patients who were not hospitalized at the initial time of presentation (discharged home from the emergency department) presented at a mean 5.1 days (SD: 4.0) into the COVID-19 disease course and none were subsequently hospitalized after discharge from the emergency department.

The demographic, clinical, and COVID-19-related characteristics of all participants, and participants stratified by hospitalization status are shown in Table 1. There was significantly higher prevalence of allergic rhinitis in hospitalized patients (p=0.007). Although the differences in prevalence did not reach statistical significance, there were more patients in the hospitalized group who were active smokers (p=0.059) and a history of heart disease (p=0.061). With respect to COVID-19 symptoms assessed, the cohort of hospitalized more frequently reported all symptoms—OD, cough, fever, shortness of breath, headache, sore throat and rhinorrhea— (p<0.05 in all cases).

**Table 1.**
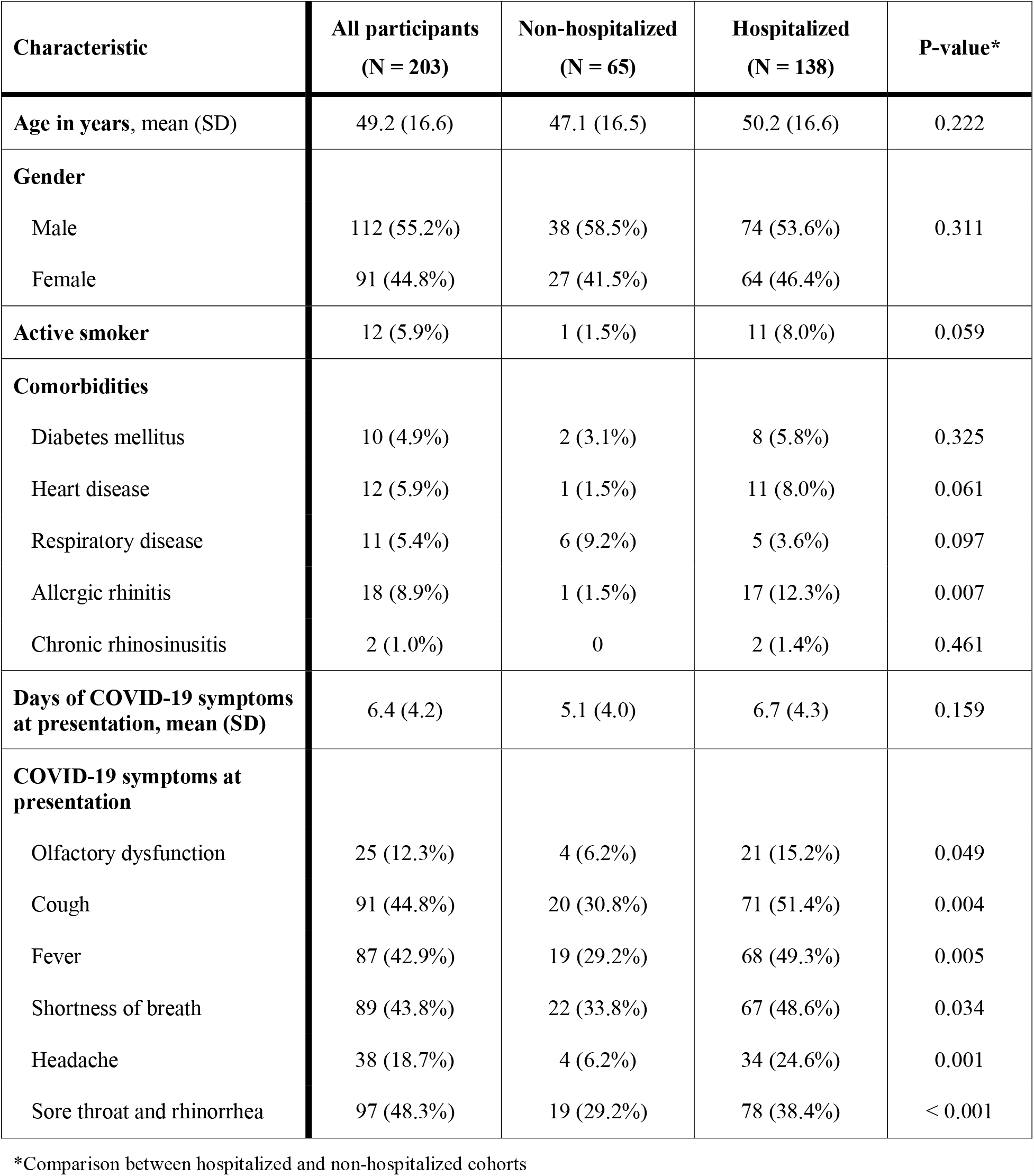
Characteristics of study participants.

The prevalence of OD at the time of presentation for the entire cohort (which was at a mean of 6.4 days after onset of COVID-19 symptoms) was 12.3%. OD was reported at the time of presentation in 15.2% of patients who were hospitalized vs. 6.2% of patients who were not hospitalized (p=0.049). Amongst patients who had OD, the mean severity of OD in hospitalized patients was 6.9 (SD: 2.6) while the mean severity of OD in non-hospitalized patients was 8.3 (SD: 5.78), which was not a statistically significant difference (p=0.508).

### Association of olfactory dysfunction of hospitalization for COVID-19

On univariate logistic regression, OD was associated with hospitalization (odds ratio [OR] = 2.47, 95%CI: 1.085 – 6.911, p = 0.049). We considered the possibility that this finding was due to differences in the length of time of COVID-19 symptoms before presentation. However, we found no significant difference in the days of COVID-19 symptoms at the time of presentation between the hospitalized group (mean: 6.7 days, SD: 4.3 days) and the non-hospitalized group (mean: 5.1 days, SD: 4.0 days) (p=0.159).

We next used a multivariable logistic regression model to check for association between hospitalization (as dependent variable) and OD (as independent variable) while controlling for age, gender, active tobacco smoking, allergic rhinitis, comorbid heart disease, comorbid respiratory disease and comorbid diabetes mellitus. We found that hospitalization was still positively associated with OD (OR=3.22, 95% CI: 0.57-18.31, p=0.188) but while the point estimate for association suggested a possible association on multivariable analysis, this was not statistically significant. We next evaluated the utility of OD as a symptom predictive of hospitalization. In our cohort, the presence of OD had 84.0% sensitivity (95%CI: 63.9% – 95.5%) but only 34.3% (95%CI: 63.9% - 41.7%) specificity for identifying patients requiring hospitalization. We also evaluated severity of OD at presentation as a predictor of hospitalization and found that severity of OD was not a statistically significant predictor of hospitalization for COVID-19 (AUC = 0.462, 95%CI: 0.251 – 0.673, p = 0.834)

## Discussion

Although initially underappreciated, OD is now a recognized symptom of COVID-19.^13, 18^ In fact, OD has proven to be a clinically informative symptom of COVID-19 in several ways. The presence of OD has been identified as a predictor of COVID-19, both in patients with flu-like symptoms as well as those who are otherwise asymptomatic.^19-22^ OD has been shown to be a typically early symptom of COVID-19, occurring in most patients within a few days of the onset of COVID-19.^7, 8^ Moreover, OD may be the sole presenting symptom of COVID-19 in up to a quarter of patients. ^9^ Consequently, OD has been recognized as an important and informative characteristic of COVID-19 with public health utility in identifying otherwise asymptomatic or pre-symptomatic COVID-19 patients who may unknowingly transmit the disease to others.^23^ Studies of OD in COVID-19 have also suggested that OD may have prognostic significance with respect to the subsequent COVID-19 disease course.^7, 14, 24^ However, studies reporting the significance of OD as a prognostic indicator of the subsequent COVID-19 disease course have so far been conflicting and therefore data from more centers are needed in this regard. Herein we report the results of a multicenter center study of OD as a predictor of hospitalization for COVID-19 in patients presenting to the emergency departments of thirteen different hospitals in the Kurdistan province of Iran. In our cohort of patients, patients who were hospitalized had a greater prevalence of OD and that OD was a sensitive (84.0%) but not specific (34.3%) predictor of hospitalization for COVID-19.

Several studies have reported that OD may be a prognostic indicator of the severity of the COVID-19 disease course. In a retrospective review of 169 patients who tested positive for COVID-19 at their institution, Yan et al reported that hospital admission was associated with an intact sense of smell while OD was associated with outpatient care.^14^ In a different study, however, Speth et al performed a cross-sectional study of patients testing positive for COVID-19 at their institution and found that the severity of OD experienced was positively correlated with the severity of shortness of breath experienced by patients, suggesting that OD was associated with a more severe COVID-19 disease course.^7^ Consistent with this finding, Benezit et al used a mobile application to study COVID-19 symptoms and found that OD was associated with hospitalization for COVID-19.^24^ Given the present dearth of evidence and conflicting reports that exist, we sought to determine whether in our patient population, OD was a predictor of hospitalization for COVID-19.

In our cohort of patients, prospectively recruited and evaluated for OD during presentation for COVID-19 at the emergency departments of 13 hospitals in the Kurdistan province of Iran, we also found a higher prevalence of OD in COVID-19 patients needing to be hospitalized compared to those who underwent outpatient management. These patients presented to these emergency departments at a mean of 6 days after the onset of COVID-19 symptoms, with no difference between those who were hospitalized compared to those who were not hospitalized for COVID-19. In our entire cohort, we found the prevalence of subjectively reported OD to be 12.3, which is a lower prevalence than the general 50 – 60% prevalence of OD that has been reported in systematic reviews of COVID-19 patients.^4^ but closer the 29% prevalence of subjectively reported OD in Iranian COVID-19 patients by Moein et al.^20^ Moreover, it is important to note that our cohort of hospitalized COVID-19 patients had a higher prevalence of all COVID-19 symptoms that we assessed than COVID-19 patients who were not hospitalized for COVID-19. Using regression analysis, we found that OD was associated with hospitalization using univariate association. However, while the point estimate for association between hospitalization and OD suggested a positive association on multivariable association, this association was not statistically significant after controlling for confounding demographic and clinical characteristics.

Our study results are consistent with the previous studies by Speth et al and Benezit et al who found OD to be associated with more severe COVID-19.^7, 24^ In our cohort of patients, those who were hospitalized also had a greater prevalence of all symptoms of COVID-19, not just OD. This is similar to findings by Speth et al who found that OD was associated with more severe COVID-19 symptoms in a Swiss cohort. ^7^ However, our results—including the finding that the statistically significant association between hospitalization and OD disappears after controlling for comorbidities—also suggest that negatively-prognostic OD (i.e. associating with more hospitalization) occurs in the setting of a greater prevalence of other COVID-19 symptoms and other risk factors for severe disease such as cardiovascular and respiratory comorbidities. Thus while OD as a single variable may be associated with COVID-19 disease severity, other underlying patient characteristics may be more directly responsible for modification of the COVID-19 disease course, which may explain why previous studies have been conflicting.

Our study should be interpreted within the constraints of its limitations. First, although this was a multicenter study, all participants were from the Kurdistan province of Iran and subjective OD may be differentially reported than in other parts of the world. All of the patients who participated in our study also were presenting to an emergency department, which suggests that these patients were experiencing more severe symptoms in general, in contrast to other studies that have reported findings for COVID-19 patients with mild to moderate symptoms. Finally, we did not use objective assessment of OD due to limitations of practicality in the setting of emergency department treatment. As reported by Moein et al, who also studied Iranian COVID-19 patients, the subjectively reported prevalence of OD may be significantly lower than OD determined by objective testing.^20^

## Conclusion

Amongst patients presenting to emergency departments for COVID-19, patient-reported OD was more prevalent in those needing hospitalization for COVID-19. However, patient-reported OD was a component of a generally higher prevalence of COVID-19 symptoms— including a greater prevalence of symptoms such as cough, fever, shortness of breath, headache, rhinorrhea and sore throat—as well as other high-risk comorbidities. Patient-reported OD in isolation may be a predictor of more severe COVID-19 and the need for hospitalization, but this association may be driven by other underlying risk factors for more severe disease.

## Data Availability

All data are available upon request.

## References

1. Guo YR, Cao QD, Hong ZS, Tan YY, Chen SD, Jin HJ, et al. The origin, transmission and clinical therapies on coronavirus disease 2019 (COVID-19) outbreak - an update on the status. Mil Med Res. 2020;7(1):11.

2. Wu YC, Chen CS, Chan YJ. The outbreak of COVID-19: An overview. Journal of the Chinese Medical Association : JCMA. 2020;83(3):217–20.

3. Borges do Nascimento IJ, Cacic N, Abdulazeem HM, von Groote TC, Jayarajah U, Weerasekara I, et al. Novel Coronavirus Infection (COVID-19) in Humans: A Scoping Review and Meta-Analysis. J Clin Med. 2020;9(4).

4. Tong JY, Wong A, Zhu D, Fastenberg JH, Tham T. The Prevalence of Olfactory and Gustatory Dysfunction in COVID-19 Patients: A Systematic Review and Meta-analysis. Otolaryngology--head and neck surgery : official journal of American Academy of Otolaryngology-Head and Neck Surgery. 2020;163(1):3–11.

5. Borsetto D, Hopkins C, Philips V, Obholzer R, Tirelli G, Polesel J, et al. Self-reported alteration of sense of smell or taste in patients with COVID-19: a systematic review and meta-analysis on 3563 patients. Rhinology. 2020.

6. Vaira LA, Salzano G, De Riu G. The importance of olfactory and gustatory disorders as early symptoms of coronavirus disease (COVID-19). Br J Oral Maxillofac Surg. 2020;58(5):615–6.

7. Speth MM, Singer-Cornelius T, Oberle M, Gengler I, Brockmeier SJ, Sedaghat AR. Olfactory Dysfunction and Sinonasal Symptomatology in COVID-19: Prevalence, Severity, Timing, and Associated Characteristics. Otolaryngology--head and neck surgery : official journal of American Academy of Otolaryngology-Head and Neck Surgery. 2020;163(1):114–20.

8. Speth MM, Singer-Cornelius T, Oberle M, Gengler I, Brockmeier SJ, Sedaghat AR. Time scale for resolution of olfactory dysfunction in COVID-19. Rhinology. 2020.

9. Kaye R, Chang CWD, Kazahaya K, Brereton J, Denneny JC, 3rd. COVID-19 Anosmia Reporting Tool: Initial Findings. Otolaryngology--head and neck surgery : official journal of American Academy of Otolaryngology-Head and Neck Surgery. 2020;163(1):132–4.

10. Lechien JR, Chiesa-Estomba CM, De Siati DR, Horoi M, Le Bon SD, Rodriguez A, et al. Olfactory and gustatory dysfunctions as a clinical presentation of mild-to-moderate forms of the coronavirus disease (COVID-19): a multicenter European study. European archives of oto-rhino-laryngology : official journal of the European Federation of Oto-Rhino-Laryngological Societies (EUFOS) : affiliated with the German Society for Oto-Rhino-Laryngology - Head and Neck Surgery. 2020;277(8):2251–61.

11. Whitcroft KL, Hummel T. Olfactory Dysfunction in COVID-19: Diagnosis and Management. Jama. 2020.

12. Hopkins C KN. Loss of sense of smell as marker of COVID-19 infection: joint statement from the British Rhinological Society and ENT-UK.: The British Rhinological Society and ENT-UK; 2020 [updated March 21, 2020. Available from: https://www.entuk.org/sites/default/files/files/Loss%20of%20sense%20of%20smell%20as%20marker%20of%20COVID.pdf.

13. American Academy of Otolaryngology - Head and Neck Surgery. Anosmia, hyposmia, and dysgeusia symptoms of coronavirus disease: American Academy of Otolaryngology-Head and Neck Surgery 2020 [Available from: https://www.entnet.org/content/aao-hns-anosmia-hyposmia-and-dysgeusia-symptoms-coronavirus-disease.

14. Yan CH, Faraji F, Prajapati DP, Ostrander BT, DeConde AS. Self-reported olfactory loss associates with outpatient clinical course in COVID-19. Int Forum Allergy Rhinol. 2020;10(7):821–31.

15. Goldman JD, Lye DCB, Hui DS, Marks KM, Bruno R, Montejano R, et al. Remdesivir for 5 or 10 Days in Patients with Severe Covid-19. The New England journal of medicine. 2020.

16. Martinez MA. Clinical trials of repurposed antivirals for SARS-CoV-2. Antimicrobial agents and chemotherapy. 2020.

17. Ministry of Health and Medical Education of Iran. [COVID-19 diagnosis and treatment flowchart in out-patient and in-patient levels]. In: Treatment DoHaDo, editor. Tehran, Iran: Ministry of Health and Medical Education of Iran; 2020.

18. ENT U. Guidance for ENT during the COVID-19 pandemic. Accessed March. 2020;23.

19. Yan CH, Faraji F, Prajapati DP, Boone CE, DeConde AS. Association of chemosensory dysfunction and COVID-19 in patients presenting with influenza-like symptoms. Int Forum Allergy Rhinol. 2020;10(7):806–13.

20. Moein ST, Hashemian SM, Mansourafshar B, Khorram-Tousi A, Tabarsi P, Doty RL. Smell dysfunction: a biomarker for COVID-19. Int Forum Allergy Rhinol. 2020.

21. Bénézit F, Le Turnier P, Declerck C, Paillé C, Revest M, Dubée V, et al. Utility of hyposmia and hypogeusia for the diagnosis of COVID-19. The Lancet Infectious diseases. 2020.

22. Salmon D, Bartier S, Hautefort C, Nguyen Y, Nevoux J, Hamel AL, et al. Self-reported loss of smell without nasal obstruction to identify COVID-19. The multicenter CORANOSMIA cohort study. The Journal of infection. 2020.

23. Sedaghat AR, Gengler I, Speth MM. Olfactory Dysfunction: A Highly Prevalent Symptom of COVID-19 With Public Health Significance. Otolaryngology--head and neck surgery : official journal of American Academy of Otolaryngology-Head and Neck Surgery. 2020;163(1):12–5.

24. Denis F, Galmiche S, Dinh A, Fontanet A, Scherpereel A, Benezit F, et al. Epidemiological Observations on the Association Between Anosmia and COVID-19 Infection: Analysis of Data From a Self-Assessment Web Application. Journal of medical Internet research. 2020;22(6):e19855.

